# Improving Real-world Antigen Test Sensitivity Estimation through Target Distribution Balancing

**DOI:** 10.1101/2024.10.25.24316137

**Authors:** Miguel Bosch, Raul Colmenares, Adriana Moreno, Jose Arocha, Sina Hoche, Auris García, Daniela Hall, Dawlyn Garcia, Lindsey Rudtner, Nol Salcedo, Irene Bosch

## Abstract

**Background:** Sensitivity is a key measure of lateral-flow antigen test (AT) performance, typically compared against qRT-PCR as the gold standard. For COVID-19, diagnostic sensitivity assesses the ability of ATs to detect SARS-CoV-2 nucleoprotein. However, sensitivity estimates can be strongly skewed by variations of the target concentrations within the clinical sample sets. Independent studies evaluating ATs from different manufacturers often display disparate target concentration distributions, making it difficult to compare sensitivity across products. We propose a new methodology to enhance the accuracy of sensitivity calculations, ensuring more reliable comparisons across ATs.

**Methods:** Sensitivity is estimated by modeling the probability of positive agreement (PPA) as a function of qRT-PCR cycle thresholds (Cts) via logistic regression of antigen test results. Raw sensitivity is calculated as the ratio of antigen test positives to total PCR positives. Adjusted sensitivity is derived by applying the PPA function to a reference concentration distribution, enabling uniform sensitivity comparisons across tests. This approach minimizes the impact of uneven sampling and external factors, as demonstrated using clinical data from a study in Chelsea, Massachusetts, USA.

**Findings:** Over two years, paired antigen and PCR positive tests from four AT suppliers were analyzed: A (211 tests), B (156), C (85), and D (43). The qRT-PCR Ct distributions varied, with suppliers A and D having more high viral load samples, while supplier C had more low viral load samples, causing significant discrepancies in raw sensitivity. Using the PPA function estimated from each supplier’s dataset, we calculated the corresponding adjusted sensitivities for common reference Ct distributions, highlighting how sample heterogeneity impacts raw sensitivity. Our approach successfully mitigates this variability, allowing for more accurate sensitivity comparisons.

**Interpretation:** This study demonstrates that sensitivity estimates from real-world data are susceptible to deviations caused by external factors, particularly the heterogeneity of qRT-PCR Ct distributions across studies. We present data supporting a novel methodology that adjusts for this variability by calculating the PPA function from raw data and determining the expected sensitivity based on a reference distribution of qRT-PCR Cts, allowing for more consistent and accurate sensitivity assessments.

**Evidence before this study:** Regulatory guidelines for antigen test (AT) performance generally require a balanced representation of low, mid, and high viral concentrations, though real-world sample distributions are highly variable. Previous studies often focus on sensitivity calculations, overlooking the impact of viral load distribution (Ct values) on results. Some studies use logistic regression to estimate the probability of positive agreement as a function of viral load, but no prior work has proposed adjusting sensitivity estimates based on a reference distribution of viral concentration.

**Added value of the Study:** This study presents a robust mathematical approach to adjust sensitivity estimates based on a standardized reference distribution of viral load, improving the precision of this key performance measure. By estimating the probability of positive agreement (PPA) as a function of viral load, we offer a more accurate assessment of AT product performance. We emphasize the importance of mitigating sample variability, showing how this method can enhance quality control and support regulatory oversight of antigen test performance.

**Implications of all the available evidence:** Our study underscores the limitations of calculating AT sensitivity directly from raw field data, which can lead to inaccurate evaluations. By applying our methodology, performance monitoring of ATs can be improved through standardized metrics, allowing for more reliable assessments. This approach helps both manufacturers and regulators establish clearer benchmarks for AT evaluation and comparison, addressing concerns about the sensitivity of antigen tests relative to gold-standard molecular methods. These improvements are critical for ensuring public confidence and regulatory accuracy in AT performance.

## Introduction

Antigen tests have been a common tool utilized to provide evidence for diagnosis and health care decisions, and have been used as such for various decades^1,2,3,4^. During the Covid-19 pandemic, the worldwide use of antigen tests demonstrated relevance for disease monitoring and diagnosis of new cases^5,6^. Antigen tests are rapid, economical, portable, can be self-administered, quick to develop, and provide direct evidence for the presence of the pathogen in the tested sample – this combination is not equaled by more sophisticated laboratory tests. Due to its increasing use, it is important to accurately evaluate the AT performance for quality and regulatory control.

The common statistic used to evaluate the positive agreement performance of antigen tests has been the sensitivity, calculated over a set of samples known to be positive by a gold standard reference. However, sensitivity of antigen tests is known to be strongly dependent on the sample viral-load.^7,8,9^ Hence, the sensitivity *perse* is not an appropriate measure of the AT positive agreement performance, because it is largely dependent on the distribution of viral load in the statistic support (i.e., the set of samples used to calculate the statistic). Instead, a description of the probability of positive agreement (PPA) as a function of the viral load is a more accurate measure of the positive agreement performance. The PPA function is commonly calculated with a logistic regression of the binary test result positive agreement (1= agreement, 0=disagreement) against a variable related with the viral load. In the case of Covid-19 disease the viral load measure is commonly the qRT-PCR cycle (Ct) result^10,11^; hence, the PPA is a function of the Cts.

Once the PPA function of a given antigen test supplier’s product has been estimated from collected data in real-world application conditions, it is straightforward to estimate the expected sensitivity for any given Ct distribution or Ct sample set. This is particularly useful for equalizing the expected sensitivity to a common standard or reference distribution of Cts for comparison of the performance across supplier’s product quality or regulatory purposes. This process removes the bias introduced into the sensitivity by the circumstantial uneven representation of viral load in the statistic support (i.e. the data used to calculate the statistic).

Common methods used to calculate the sensitivity of an AT product, whether for regulatory compliance or communication purposes, typically involve collecting real-world test results paired with the qRT-PCR gold standard. However, in the case of lateral-flow antigen tests, sensitivity uncertainty does not adhere to a straightforward Bernoulli process, as the underlying positive agreement probability is not constant and is instead conditioned by the viral load. To accurately calculate sensitivity, it would be necessary to segment the collected samples based on the most influential variable affecting the underlying probability, using a standard reference histogram^12^. In our scenario, the influential variable is the viral load, measured as Ct. Yet, implementing such a process in the field with real-world data would be cumbersome and would require a much larger dataset. The proposed balancing method overcomes this challenge by adjusting the raw sensitivity calculation to any desired standardized reference distribution of the viral load, without the need of extended data collection and segmentation.

## Methods

### Study description

We conducted a study of AT use in real-world conditions in the city of Chelsea, Massachusetts during years 2022-2023 (Advarra Protocol Number 00059157). The objectives of the study were multifold: (1) performing frequent Covid-19 testing at two vulnerable population sites (elderly housing), (2) evaluating the performance of ATs from different suppliers in the laboratory and in the real-world context, (3) collecting longitudinal (time series) AT data for qRT-PCR positive samples, and (4) implementing a digital AT data collection platform.

The participants of the study were enrolled after consent. The participant was provided with ATs for self-testing, a single AT of any of the available four participating suppliers. The tests were Home tests, and data was self-logged by the participants into an internet based informatic platform. The participants registered the AT results (their own assessment) and uploaded a photograph of the test after completion (15 min). The fraction of positive AT tests was followed daily with paired qRT-PCR testing. A random number of negative AT tests were also analyzed by PCR.

The data analyzed come from ATs provided by four different suppliers, labeled A through D, and the corresponding qRT-PCR test results, all of which were processed in the same CLIA-certified laboratory. The total negative qRT-PCR tests were 57 for A, 91 for B, 145 for C and 114 for D and positive qRT-PCR tests were 211 for A, 156 for B, 84 for C and 43 for D. Each participant was tested with a single brand (i.e., AT supplier), so the distribution of viral load could be different between the data collected for each brand. These data were used to calculate the AT performance statistics and demonstrate the methodology for balancing the sensitivity according to a reference standard distribution.

### Probability of positive agreement function

As commonly used, AT operative reading involves steps of device reaction to the sample, waiting time, observation and interpretation of the result by the user. Although various device designs are used (e.g., cassette, card), the user will see the presence or absence of color in a marked zone (*test band*) on a nitrocellulose strip used for the lateral-flow reaction process. The result is considered *positive* when the test band can be distinguished from the background even if the signal is faint versus a *negative* if the test band cannot be seen visually by the user. For performance statistics, the result of the AT provided by the user is compared to the gold standard reference test, which in the case of this study was Covid-19 qRT-PCR.

For the positive agreement analysis of each AT supplier dataset, we compare only the AT results that have a paired positive qRT-PCR result (i.e., having a positive qRT-PCR result in a swab sample taken the same day as the antigen test swab sample). For the logistic regression analysis, we identify the AT results with a binary variable: 1 for a positive results (agreement with the standard test), and 0 for a negative result (disagreement with the standard test). The outcome of the user assessment can be described by a binary random variable. We model the PPA with a logistic function, having the qRT-PCR cycle count (Ct) as the function domain (i.e. the independent variable). Logistic regression is well-known analysis to estimate the probability as a function of a dependent variable^13^. It has been used to describe the probability of positive agreement in antigen tests^14^.

In addition to estimating the PPA function that characterized each AT supplier data, the regression also accounts the uncertainties of the probability function and parameters. We implement the logistic regression with a Bayesian approach, combining the objectives of (1) fitting the binary observed data, and (2) honoring the Clopper-Pearson binomial confidence limits at the raw Ct data sensitivity prediction. Hence, the posterior model uncertainty description ensures compliance with the Clopper-Pearson confidence limits for the sensitivity at the raw Ct data. The numerical calculations are performed by Markov Chain Monte Carlo methods.

### Sensitivity balancing method

The sensitivity, *s*, is the fraction of the positive agreement cases divided by the total positive cases in the experimental gold standard results (e.g., collected real-world AT binary data on positive qRT-PCR cases). Based on the PPA function characterized for each AT supplier data, we can estimate the sensitivity for any set of Ct cases. Let’s consider that the experimental data for a given AT supplier involves *N* cases with qRT-PCR cycle counts, **x** = {*x*_1_, *x*_2_, … *x*_*n*_, …, *x*_*N*_ }. The expected value of the sensitivity is the average of the PPA function, *p*(*x*), over the cases,

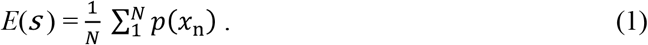

Likewise, the expected sensitivity over a data support with any Ct probability density function (PDF), *g*(*x*), is given by the probability product integration,

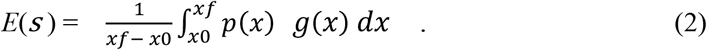

The equations (1) and (2) use the PPA function to calculate the expected sensitivity over a specific Ct data or distribution. Adequate comparison of sensitivity across different AT datasets requires a transformation of the raw sensitivity (i.e., calculated from the observed data) to the expected sensitivity over a common reference of Ct data values, or a Ct support distribution (i.e. histogram or PDF). Considering observed binary data for various AT suppliers, our proposed process to equalize the sensitivity support involves:

a. Estimating the PPA functions by logistic regression of the observed AT binary data for each one of the supplier’s datasets,
b. defining a common reference Ct distribution, by a PDF *g*(*x*) and,
c. calculating for each AT supplier dataset the estimated sensitivity over the common reference Ct support by equation (2).

The integral (2) can be evaluated by the domain discretization. Alternatively, it can be calculated by Monte Carlo integration, pulling a set of Ct realizations from *g*(*x*) and using expression (1). Balancing the viral load support could also take as reference the sample set of one of the AT suppliers, e.g., supplier A. Then, the estimated sensitivity of the other suppliers can be estimated over the same Ct support of supplier A data for appropriate comparison. In the latter case the method follows:

a. Estimating the PPA functions by logistic regression of the observed AT binary data for each one of the other supplier’s datasets and,
b. calculating the expected sensitivity for each one of the supplier’s datasets over the reference Ct collection of supplier A using expression (1).

The described viral load balance processes remove the effect of the Ct field data distribution on the sensitivity, providing a common base for comparison and evaluation of the positive agreement performance.

## Results

### Raw positive agreement statistics

This section describes the basic performance statistics of the ATs of the four suppliers analyzed, and the estimated PPA functions, based on the binary data collected from the Chelsea study. The agreement matrix was determined for each supplier AT, and the common performance agreement fractions were calculated: sensitivity, specificity, positive prediction, negative prediction and total prediction. Table 1 displays the basic performance statistics for each one of the test suppliers, including the Clopper-Pearson 95% confidence limits^15^.

**Table 1.**
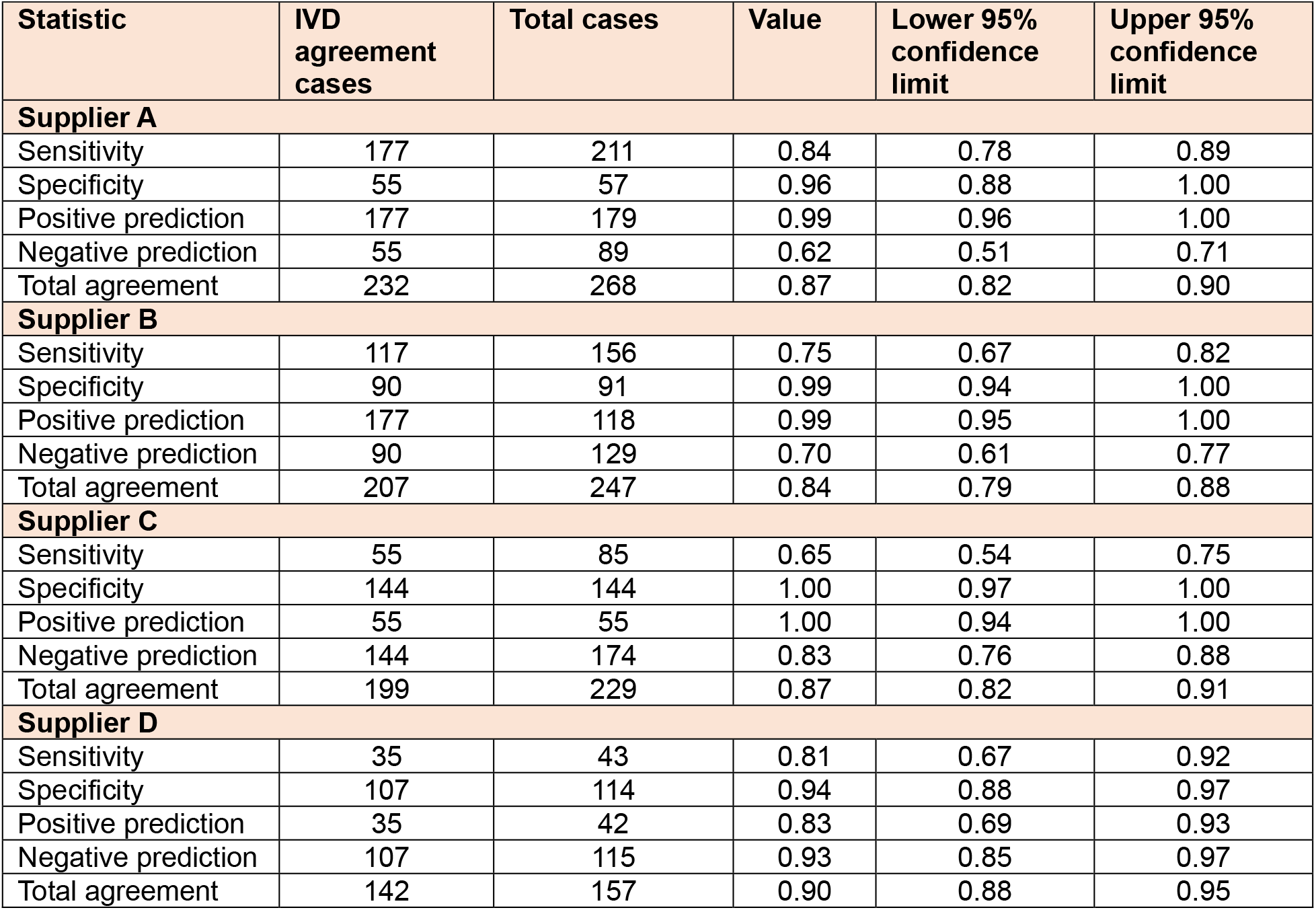
Basic performance statistics of IVD suppliers A, B, C and D.

| Table 1. Basic performance statistics of IVD suppliers A, B, C and D |  |  |  |  |  |
| --- | --- | --- | --- | --- | --- |
| Statistic | IVD agreement cases | Total cases | Value | Lower 95% confidence limit | Upper 95% confidence limit |
| **Supplier A** |  |  |  |  |  |
| Sensitivity | 177 | 211 | 0.84 | 0.78 | 0.89 |
| Specificity | 55 | 57 | 0.96 | 0.88 | 1.00 |
| Positive prediction | 177 | 179 | 0.99 | 0.96 | 1.00 |
| Negative prediction | 55 | 89 | 0.62 | 0.51 | 0.71 |
| Total agreement | 232 | 268 | 0.87 | 0.82 | 0.90 |
| **Supplier B** |  |  |  |  |  |
| Sensitivity | 117 | 156 | 0.75 | 0.67 | 0.82 |
| Specificity | 90 | 91 | 0.99 | 0.94 | 1.00 |
| Positive prediction | 177 | 118 | 0.99 | 0.95 | 1.00 |
| Negative prediction | 90 | 129 | 0.70 | 0.61 | 0.77 |
| Total agreement | 207 | 247 | 0.84 | 0.79 | 0.88 |
| **Supplier C** |  |  |  |  |  |
| Sensitivity | 55 | 85 | 0.65 | 0.54 | 0.75 |
| Specificity | 144 | 144 | 1.00 | 0.97 | 1.00 |
| Positive prediction | 55 | 55 | 1.00 | 0.94 | 1.00 |
| Negative prediction | 144 | 174 | 0.83 | 0.76 | 0.88 |
| Total agreement | 199 | 229 | 0.87 | 0.82 | 0.91 |
| **Supplier D** |  |  |  |  |  |
| Sensitivity | 35 | 43 | 0.81 | 0.67 | 0.92 |
| Specificity | 107 | 114 | 0.94 | 0.88 | 0.97 |
| Positive prediction | 35 | 42 | 0.83 | 0.69 | 0.93 |
| Negative prediction | 107 | 115 | 0.93 | 0.85 | 0.97 |
| Total agreement | 142 | 157 | 0.90 | 0.88 | 0.95 |

Sensitivities show large differences across the suppliers A, B, C and D, with values 0·84, 0·75, 0·65 and 0·81 correspondingly. A comparison plot of the raw AT sensitivities for each supplier and confidence limits is shown in figure 1. Differences are significant with a large departure of 19% (percentual points of the sensitivity) between suppliers A and C. However, figure 2 shows that the histograms of qRT-PCR Cts supporting the sensitivity calculations have marked differences across the suppliers. Note that suppliers A and D have a larger proportion of low Cts (large viral sample concentration). On the other hand, supplier C has a larger representation of large Cts (low viral sample concentration).

**Figure 1.**
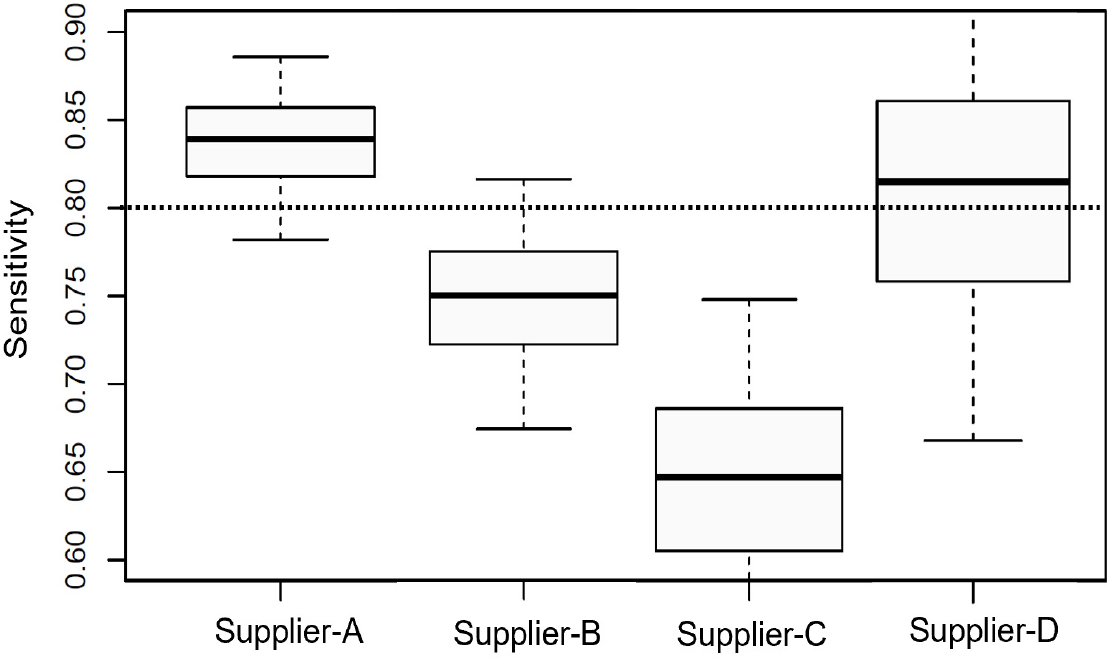
Sensitivities calculated from the raw data from the Chelsea study for suppliers A, B, C and D. The boxes show 50% and the segments 95% confidence limits calculated with the Clopper-Pearson method. The dotted line shows the 0·8 sensitivity shred hold.

**Figure 2.**
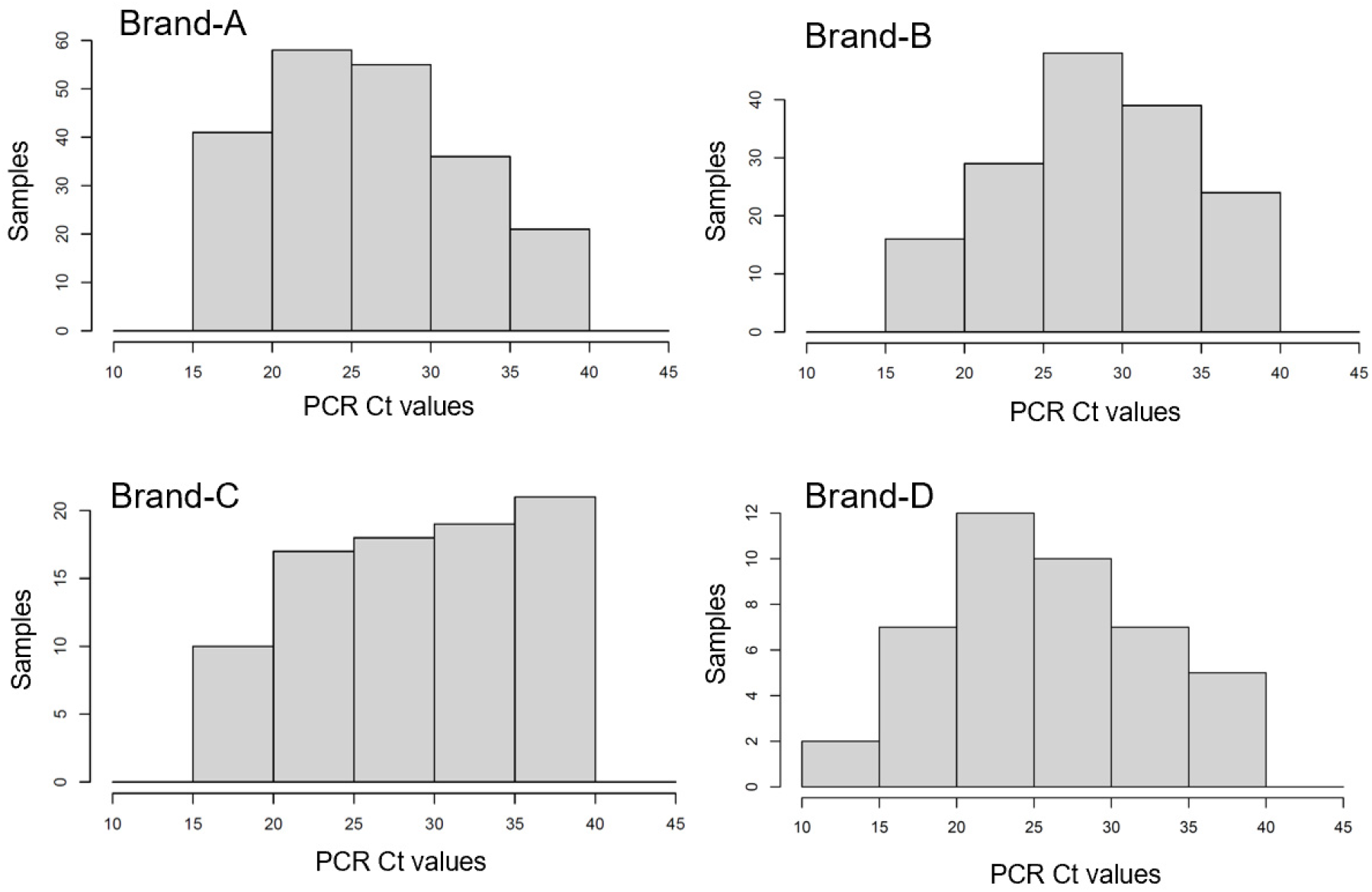
Histograms of the samples qRT-PCR cycle count (Cts) for the four datasets analyzed, each one corresponding to a different AT supplier. The Cts shown are the average between the cycle counts of gene N and gene ORFab.

### Probability of positive agreement functions

Raw sensitivities, as shown in figure 1, superpose the effects of the viral concentration support to the true performance of the ATs. In what proportion the differences in sensitivity shown in the figure is caused by the uneven distribution of viral concentration shown in figure 2? Following our method, a first step to decouple the two effects is estimating the PPA function from the raw data of each AT supplier by logistic regression, as explained in the previous section. Figure 3 shows the binary data collected for each AT supplier plotted against the Cts, and the estimated PPA function for each test supplier. The estimation of the PPA function by fitting the observed binary AT data also provides description of the uncertainties in the PPA function. This is illustrated in figure 3 by the 95% confidence intervals of the PPA function; our formulation estimates the full distribution of the PPA conditioned to the Ct value.

**Figure 3.**
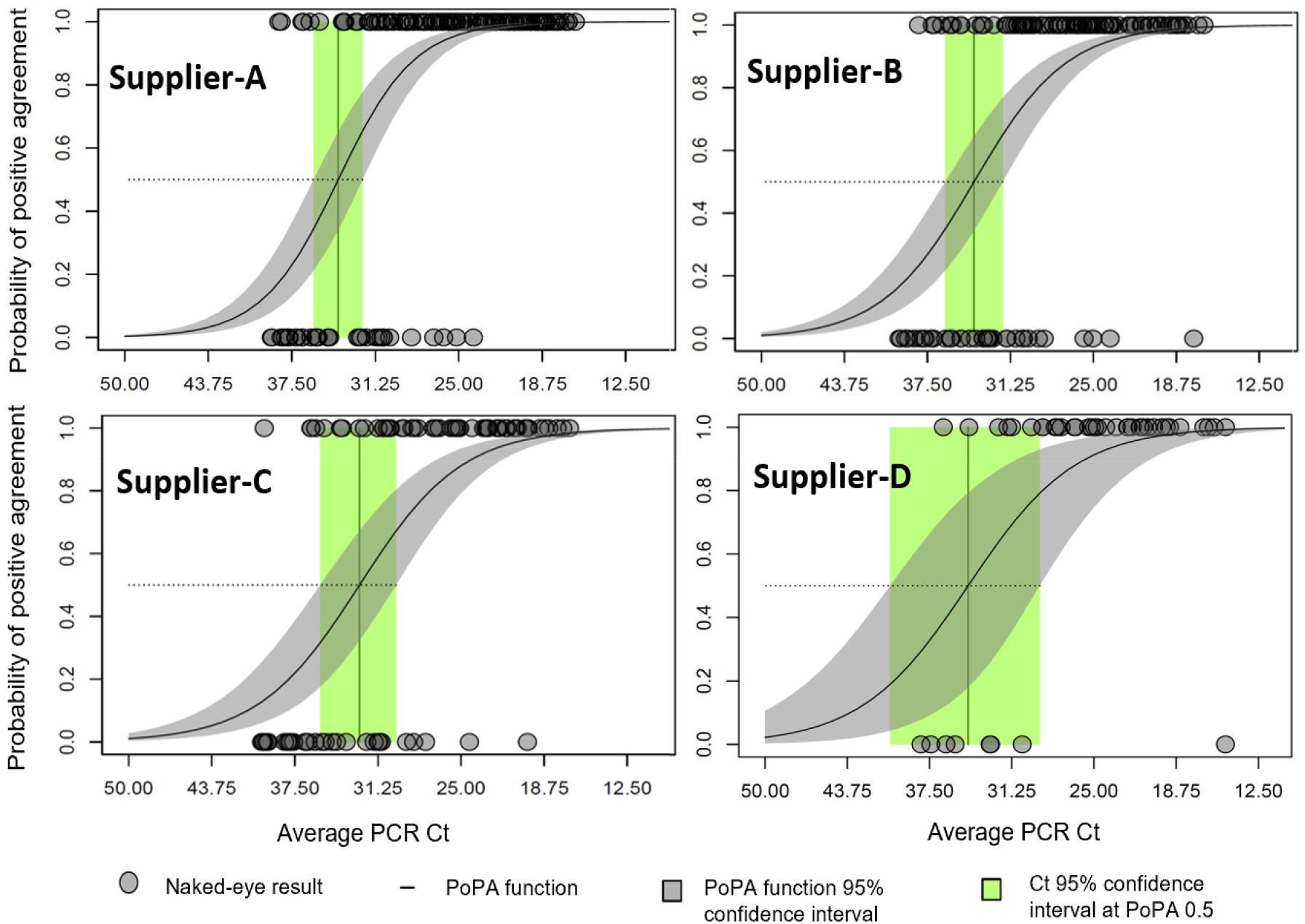
Probability of positive agreement (PPA) as function of the qRT-PCR cycle count (Ct) for naked-eye assessments of the Chelsea project participants after self-application of the antigen tests. Naked-eye assessments of the antigen test result are plotted in the vertical axis with value 1 for positive and 0 for negative. The PPA function is obtained by logistic regression of the binary naked-eye results, and shows the strong dependency of the agreement probability with the qRT-PCR Cts.

Figure 3 shows that the PPA functions for suppliers A and B are similar: note that the Ct of median probability (i.e., limit of detection at probability p=0·5) are very close. Also, the inclination of the function is similar, slightly higher for supplier A. The PPA function for supplier C shows a slightly larger Ct at median probability and lower inclination of the function. The PPA function for supplier D is also close to the A and B functions, but showing larger uncertainties as expected form the smaller data support of supplier D.

### Reference distributions of viral-load

Due to the dependency of the sensitivity on the Ct value distribution the comparison of raw sensitivities is biased by the uneven distribution of the Ct support, as shown by the histograms in figure 2. Utilizing our support balance methodology, we computed the sensitivities of the ATs from the four suppliers across four distinct reference distributions of the Cts. The sample statistics of the PPA exhibit particular sensitivity to low positive samples, i.e., those with low viral concentration. Consequently, we opt for a uniform distribution of qRT-PCR cycles spanning 10-35 Cts, with variable proportions within the 35-40 range, to underscore the significance of representing low-positive cases in the overall sample PPA. Additionally, we employed the combined Ct distribution of all four tests as a reference, i.e., the joint positive qRT-PCR Ct counts of the four supplier’s data. The corresponding histograms for these distributions are depicted in Figure 4.

**Figure 4.**
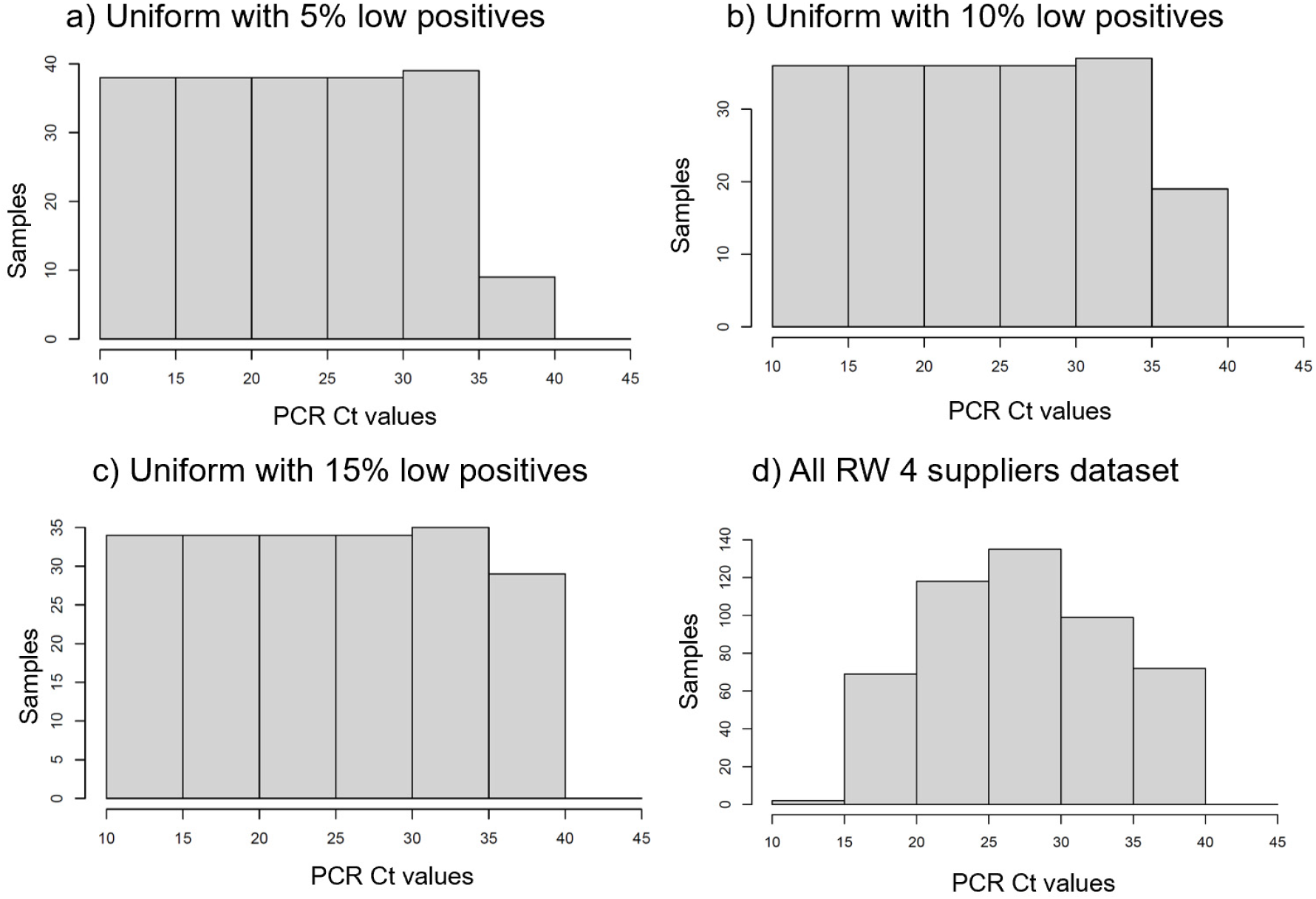
Four different reference distributions used to correct the raw data sensitivity calculations from uneven viral concentration sampling. Histograms a, b, and c correspond to 200 cases with variable fraction of low positives (35-40 Cts) and uniform distribution in the 10-35 Ct range. Histogram d corresponds to the joint four supplier’s observed data.

### Sensitivities for the reference distributions

Figure 5 shows the estimated sensitivities of the ATs of the four suppliers over each one of the reference distributions shown in figure 4, according to the described viral load balance method. It is useful to compare these results to the raw sensitivities shown in figure 1. Although the order of performance of the four suppliers has been preserved, the sensitivity differences are much smaller once removed the effect of the source support by the balancing process. The difference between suppliers A and C are only 5% points (precent of sensitivity) instead of the 19% points for the raw sensitivity calculation. Large proportion of the raw sensitivity difference between these two suppliers was caused by the over representation of large viral concentrations in supplier A samples, and over representation of low viral concentrations in supplier C samples.

**Figure 5.**
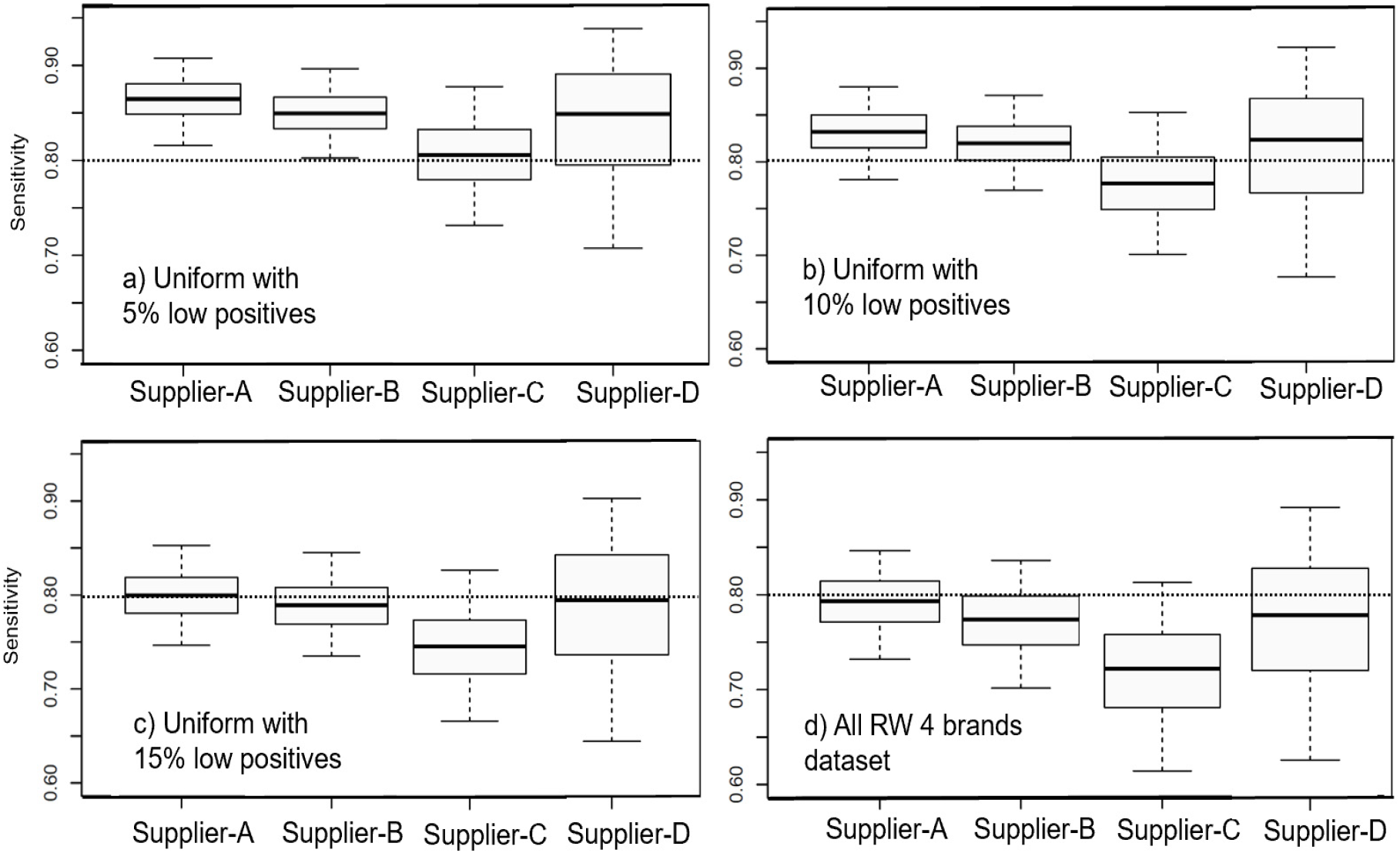
Sensitivity corrected from the uneven distribution of viral concentration across the four supplier’s datasets, and corresponding to the four optional referential distributions of qRT-PCR cycles shown in figure 4. The boxes show 50% and the segments 95% confidence limits. The dotted line shows the 0·8 sensitivity shred hold.

A second point shown in figure 5 is the strong influence of the reference distribution of Cts on the absolute values of the sensitivity. The absolute value of the sensitivity experiences important variation of the order of 8% points across the different reference distributions– larger than the difference across the supplier sensitivities. In particular, the fraction of low positives plays an important role, as expected: the PPA functions in figure 3 show that the probability of positive agreement is in all suppliers is very low for low positives (35-40 Cts). The indicated line at 0·8 sensitivity in figure 5 helps to illustrate this point. With the reference distribution including 5% of low positives the four suppliers have sensitivities over the 0·8 threshold. With the reference distribution including 15% of low positives suppliers B and C are below the threshold, whereas suppliers A and D are borderline. With the distribution that combines the four suppliers observed samples, all the suppliers are below the 0·8 threshold.

## Discussion

Due to inherent factors such as the number of participants, demographics, and sampling conditions in real-world data, the qRT-PCR Ct distribution used for sensitivity or PPA calculation may yield varying results. Due to limitations on the considerable list factors, it is feasible to take into consideration all incoming datapoints to shape a particular Ct distribution if a probability function (PPA) is applied. This practical challenge arising from the intrinsic variability of a study, including divergent viral load distributions, can be addressed through a data balancing procedure. This procedure involves estimating the PPA function, followed by recalculating sensitivity over a selected referential or standard Ct distribution. By doing so, the process mitigates the impact of uneven Ct distributions in the raw data and provides a mathematical computation of the expected PPA.

Regulatory agencies have considered incorporating low levels of the target in antigen test performance analysis to ensure less biased statistics and a comprehensive representation of both low and high viral loads. While this approach is practical, it lacks a generalizable method, focusing solely on low positive bins and neglecting to describe the remaining data ranges. This method provides an alternative for making assumptions when comparing two datasets, as it models the entire dataset, including both low and high Ct values, to achieve balanced data distribution. By modeling the distributions of the data, we encompass all outcome ranges, allowing for meaningful comparisons between groups. The statistics we propose rely on distributions rather than specific ranges.

The described methodology enhances the reliability of performance metrics for antigen tests (ATs), addressing the significant bias inherent in raw data sensitivity calculations. We propose the calculation of the PPA function characterizing the AT supplier data, whereas in the domain of Cts, copies per mL, or other sample viral concentration related variable. However, comparing supplier’s AT performance based on direct examination of the PPA function is not straightforward, as various characteristics of the PPA function (such as Ct at median probability, slope, uncertainties, etc.) have a coupled effect in the overall performance. Sensitivity offers the advantage of providing a single numerical value for evaluation and comparison. Thus, we propose the viral load adjusting methodology: defining a standard reference Ct distribution and calculating sensitivity over the reference distribution as a accurate measurement of AT positive agreement performance.

The real-world data includes factors that will bias the result of the sensitivity of the antigen test and does not reflect performance with accuracy. The method we describe allow for the adjusted and more accurate representation of the tests’ sensitivity. The model proposed provides a robust statistical analysis based on logistic regression of the positive agreement probability variable.

The raw sensitivity directly calculated from real world datasets show to be strongly influenced by the distribution of the virus concentration at the time of testing the participats of the study. Therefore, having a way to balance to a common distribution is a useful. We provide a model to adjust the raw sensitivities.

## Data Availability

All data is available at the Rapid Acceleration of Diagnostics - Underserved Populations (RADxUP, a program funded by the National Institutes of Health (NIH), under the repository conditions.

https://idx20.us

## Contributors

MB and IB contributed with the conceptualization of the methodology and manuscript writing. MB developed the mathematical formulations, algorithms, calculations and data analysis. IB designed and implemented the Chelsea data collection study. RC, DG, LR and NS contributed with data analysis and AM implemented a web-app interphase for the tool. SH, DH and AG contributed with the data collection and executed clinical study and on-site acquisition.

## Data sharing

The data are publicly available at Clinicaltrials.gov NCT05884515. Upon request at RADxUP https://myhome.radx-up.org for CorDx data.

## Acknowledgments

We thank the sponsoring of the Reagan-Udall Foundation for the FDA and to the *Rapid Acceleration of Diagnostics for Underserved Populations (RADxUP) Coordination and Data Collection Center* for the support and The Broad Institute for CorDx test collection and processing. We thank the Chelsea Housing Authority to offer guidance and a support to the study site and we thank the participants of the study for their valuable contribution and attendance. To Adriana Mendoza for coordination of the studies and to Anna Roberts at Eco Laboratory, Acton, MA, for PCR processing of the samples.

